# Arachnoid granulations may be protective against the development of shunt dependent chronic hydrocephalus after aneurysm subarachnoid hemorrhage

**DOI:** 10.1101/2021.12.25.21268402

**Authors:** Khaled Almohaimede, Fulvio Zaccagna, Ashish Kumar, Leodante da Costa, Erin Wong, Chris Heyn, Anish Kapadia

**Author notes:** Corresponding Author: Anish Kapadia.

## Abstract

**Background and Purpose:** Chronic hydrocephalus may develop as a sequela of aneurysmal subarachnoid hemorrhage, requiring long-term cerebrospinal fluid shunting. Several clinical predictors of chronic hydrocephalus and shunt dependence have been proposed. However, no anatomical predictors have been identified.

**Materials and Methods:** A retrospective cohort study was performed including 61 patients with aneurysmal subarachnoid hemorrhage. Clinical characteristics were noted for each patient including presentation World Federation of Neurosurgical Societies grade, modified Fischer grade, aneurysm characteristics, requirement for acute and chronic cerebrospinal fluid diversion, and 3-month modified Rankin scale. CT images were evaluated to determine the Evans’ index and to enumerate the number of arachnoid granulations. Association between the clinical characteristics with ventriculoperitoneal shunt insertion and the 3-month modified Rankin scale were assessed.

**Results:** The initial Evans’ index was positively associated with mFisher grade and age, but not the number of arachnoid granulations. 16.4 % patients required insertion of a ventriculoperitoneal shunt. The number of arachnoid granulations were a significant negative predictor of ventriculoperitoneal shunt insertion [OR: 0.251 (95% CI:0.073-0.862; P=0.028)]. There was significant difference in the number of arachnoid granulations between those with and without ventriculoperitoneal shunt (p=0.002). No patient with greater than 4 arachnoid granulations required a ventriculoperitoneal shunt, irrespective of severity of initial grade.

**Conclusion:** Arachnoid granulations may be protective against the development of shunt dependent chronic hydrocephalus after aneurysmal subarachnoid hemorrhage. This is irrespective of presenting hemorrhage severity. This is a potentially novel radiologic biomarker and anatomic predictor of shunt dependence.

## Introduction

Communicating hydrocephalus is defined as ventricular dilatation due to abnormal accumulation of cerebrospinal fluid (CSF) secondary to obstruction distal to the ventricular system^1^. Impairment of the usual drainage pathways via arachnoid granulations (AG) and the skull-base lymphatics due to blood products, such as in subarachnoid hemorrhage (SAH), can result in communicating hydrocephalus^2^. The incidence of hydrocephalus is estimated to be approximately 20-30% in patients presenting with aneurysmal SAH (aSAH), and has been observed at various stages during the course of the illness^3^. Resultant increase in intracranial pressure can result in neurological deterioration and even death if untreated. Temporary shunting with extra-ventricular drain (EVD) is usually necessary in case of uncompensated acute hydrocephalus. However, some patients develop chronic hydrocephalus requiring long-term CSF diversion^4^.

AG are sac-like protuberances composed of endothelial cells which protrude into venous sinuses, commonly found near the superior sagittal, straight, transverse, and sigmoid sinuses^5^, and act as a resorption conduit and allow for CSF egress from the subarachnoid space into the systemic circulation^6^. Until recently, they were thought to be the exclusive pathway for CSF egress. Recently discovered lymphatic pathways, draining via skull-base perivascular and perineural pathways, are increasingly recognized as important routes of CSF egress^7^. SAH may result in obstruction of CSF drainage via both drainage pathways. The small perineural and perivascular drainage channels can get obstructed by blood products^8^. Blood products may also directly obstruct drainage via AGs, or the ensuing inflammatory reaction from the presence of blood products can result in fibrosis resulting in decreased CSF resorption^9^.

Several factors have been proposed as clinical predictors of hydrocephalus and/or shunt dependence; however, many of these remain controversial due to conflicting evidence^10^. The presence of intraventricular hemorrhage, Hunt and Hess grade (and Fisher grade), re-hemorrhage, higher age, and infection have all been variably linked to increased risk of shunt dependence^11^. However, there is no anatomic factor that has been identified as a potential biomarker for the development of hydrocephalus and subsequent shunt dependence. Since AG are an important, and potentially the primary route for CSF egress, we hypothesise that a higher number of AGs may be protective factor for the development of hydrocephalus and shunt dependence. Hence, the aim of this study was to explore a potential correlation between the number of AGs and the development of shunt dependence in a cohort of patients with aSAH.

## Materials and Methods

### Patients’ selection

A retrospective cohort study was conducted under local REB approval at our institution. All cases of aSAH between January 1, 2015 and June 31, 2020 were initially obtained from a departmental imaging database using a keyword search of the CT reports (including the following keywords: subarachnoid hemorrhage, aneurysm). The reports and images were then reviewed and retrieved. All patients > 18 year-old, with a presentation of acute aSAH, and baseline CTA were included. Patients with poor contrast opacification of the venous system, hydrocephalus secondary to other etiology (e.g. obstructing mass), and those without imaging follow up were excluded.

A retrospective review of the electronic medical record was performed. Patient demographics, clinical status at presentation including World Federation of Neurosurgical Societies grading system (WFNS), aneurysm location, aneurysm size, requirement for acute CSF shunting with an EVD, requirement for chronic CSF shunting as indicated by insertion of a ventriculoperitoneal shunt (VPS), in-hospital complications, and 3-month modified Rankin score (mRS) were recorded.

### Image acquisition and analysis

CT scans were acquired as part of the clinical workup as per standard-of-care at our institution. CT acquisitions were performed on 64-section CT scanners (LightSpeed Plus and VCT; GE Healthcare, Milwaukee, Wisconsin). Unenhanced CT examination was performed from the skull base to the vertex with the following parameters: 120 kV(peak), 340 mA, 4 × 5 mm collimation, 1 s/rotation, and a table speed of 15 mm/rotation. CTA acquisitions were performed from C6 to the vertex in the helical half-scan mode with the following parameters: 0.7 mL/kg of iodinated contrast (to a maximum of 90 mL through an antecubital vein via at least an 18- or 20-ga angiocatheter), 120 kV(p), 270 mA, 1 s/rotation, 1.25-mm section thickness at 0.625-mm intervals, and table speed of 3.75 mm/rotation. The initial unenhanced CT images were evaluated to determine the modified Fisher (mFisher) grade and the Evans’ index as previously described^9,10^. AGs were identified as eccentric spherical low-density filling defects within the dural venous sinuses using multiplanar reformat of baseline CTA images. Examples of AGs are provided in Figure 1. The number of macroscopic AG (≥ 1mm) seen along the superior sagittal, straight, transverse, and sigmoid sinuses were assessed and enumerated.

**Figure 1:**
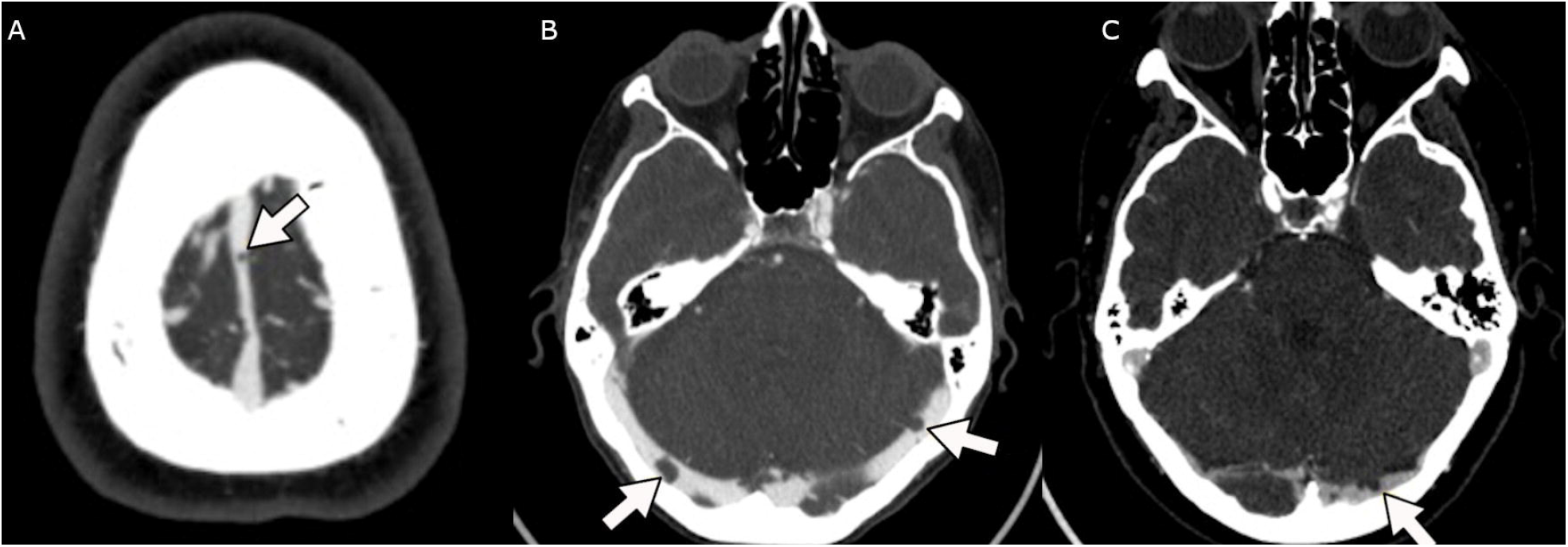
CTA images with adequate venous opacification demonstrating macroscopic arachnoid granulations (>1mm), spherical hypoattenuating filling defects within the dural venous sinuses, identified with small arrows on panel A-C.

### Statistical Analysis

Statistical analysis was performed using RStudio (version X64 3.4.4 for Windows; RStudio Inc., Boston, MA, USA, The R Foundation for Statistical Computing Platform)^12^ and JASP (version 0.16 for Windows, JASP team, Netherlands)^13^. Descriptive statistics for patients’ demographic data were generated. Normality assessment was performed using the Shapiro-Wilk test. Association between two variables was assessed using Spearman-rank correlation or Pearson correlation as appropriate. The mRS was dichotomized into groups 0-1 and 2-6 as previously described^14^. Logistic regression analysis was performed for the dichotomized mRS and the insertion of a VPS accounting for relevant demographic and clinical covariates. The Holm-Bonferroni correction was performed to control for multiple comparisons. Statistical significance was set at (p<.05).

## Results

A total of 61 patients with aSAH were included with an average age of 58.8 ± 13.1 years (29-83) comprised of 45 (74%) female (Table 1). VPS insertion was required in 16.4 % (10) of patients, consistent with rates previously reported^11^. The distribution of aneurysm location and the size of the aneurysms are presented in Table 1, along with Evan’s index on the baseline CT, mFisher grade, and WFNS. The distribution of 3-month mRS is also presented in Table 1. The baseline Evans’ index was positively associated with the mFisher grade (r=.54, p<.001) and age (r=.44, p=.003), but not the number of AGs or WFNS. The presenting WFNS was correlated with the mFisher grade (r=.41, p=.008). Amongst patients with mFisher grade 4 hemorrhage, the number of AG was correlated inversely with Evans’ index at baseline (r=-.31, p=.028) and the WFNS grade (r=-.27, p=.045).

**Table 1:** Demographic data, aneurysm location, aneurysm size, Evans’ index on initial CT, presenting World Federation of Neurosurgical Societies (WFNS) grade, modified Fisher grade, 3 month modified Rankin scale and ventriculoperotneal shunt (VPS) status.

|  |  |
| --- | --- |
| **Age** | 58.8 (+/- 13.1) |
| **Gender** | 45 (74%) Female |
| **Aneurysm Location:** |  |
| • Acom | 22 |
| • Pcom | 7 |
| • MCA | 18 |
| • ICA | 7 |
| • Basilar | 2 |
| • Anterior Choroidal ( AChA) | 1 |
| • Others | 4 |
| **Aneurysm size** | 5.4 +/- 2.6 mm |
| **Evans' Index (initial CT)** | 0.28 +/- 0.04 |
| **WFNS grade:** |  |
| • 1 | 24 (39%) |
| • 2 | 18 (30%) |
| • 3 | 2 (3%) |
| • 4 | 11 (18%) |
| • 5 | 6 (10%) |
| **Modified Fisher grade:** |  |
| • 1 | 7 (11%) |
| • 2 | 4 (7%) |
| • 3 | 10 (16%) |
| • 4 | 40 (66%) |
| **3 Months modified Rankin Scale:** |  |
| • 0 | 20 (34%) |
| • 1 | 18 (31%) |
| • 2 | 3 (5%) |
| • 3 | 4 (7%) |
| • 4 | 7 (12%) |
| • 5 | 2 (3%) |
| • 6 | 4 (7%) |
| **VPS insertion:** |  |
| • Yes | 10 (16.4 %) |
| • No | 51 (83.6 %) |

Logistic regression analysis was performed for shunt dependence as measured by VPS insertion. The number of AG was a significant negative predictor of VPS insertion when accounting for covariates including age, baseline Evans’ index, and presenting WFNS grade [OR: 0.251 (95%CI: 0.073-0.862; p=.028)] or mFisher [OR: 0.418 (95%CI: 0.198-0.881; p=.022)]. There was a significant difference in the number of AGs between patients who required and patients who did not require VPS (p=.002; Figure 2), and this was also true amongst the mFisher 4 subgroup (p=.005). No patient with 4 or more AG required a VPS, including those with mFisher grade 4 SAH. The baseline Evans’ was the only positive predictor of VPS insertion (p=.036). Logistic regression was performed to assess poor 3-month outcome (mRS 2-6); WFNS grade 5 at presentation was the only significant predictor (OR: 14.74 (1.18-184.6; p=.04), however, a weak negative trend was noted with the number of AG.

**Figure 2:**
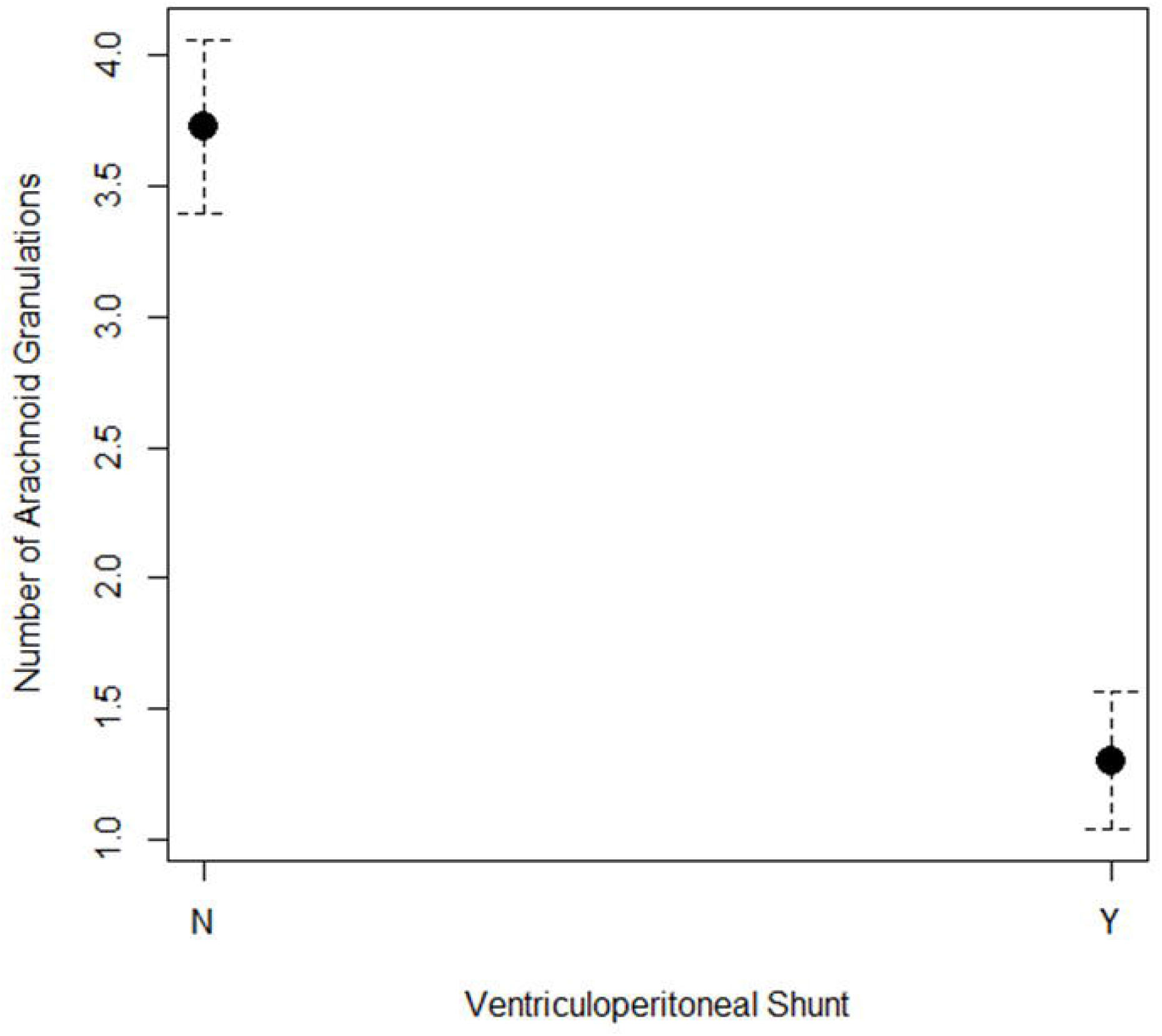
Mean number of arachnoid granulations for ventriculoperitoneal shunt (VPS) status. N: No VPS; Y: Yes VPS.

## Discussion

The current study is the first to identify the number of AGs as a potential predictor of shunt dependence in patients with aSAH. Even amongst those patients with the most severe aSAH, as classified by the mFisher grade 4, those requiring long-term CSF diversion have significantly fewer AGs. In our cohort, no patient with 4 or more AGs required a VPS irrespective of hemorrhage severity. This is a potentially novel radiologic biomarker that can be used to predict which patients may ultimately require long-term CSF diversion. These findings also raise more fundamental questions about CSF clearance.

The factors that determine the requirement for long-term CSF diversion have been extensively studied. A host of clinical factors have been variably linked to the development of shunt dependence; however, there is significant heterogeneity in the published evidence. Age ≥50 years, female gender, high Hunt-Hess grade, Glasgow Coma Scale ≤8, Fisher grade ≥3, acute hydrocephalus, external ventricular drainage (EVD) insertion, intraventricular hemorrhage, posterior circulation aneurysm, anterior communicating artery aneurysm, meningitis, and re-bleeding have all been found to be associated with shunt dependence^15^. Factors such as Fisher grade which may be linked intuitively with hydrocephalus and shunt dependence, in reality, have conflicting evidence^10^, including findings from this study. Clinical factors which were once thought to be associated with shunt dependence, such as the duration of EVD have not been linked with definite evidence^16^ The current findings are the first to provide an anatomic predictor of shunt dependence, and specifically the protective effect of AGs.

The mechanisms by which CSF egress occurs from the brain remains a topic of intense research and has come under further scrutiny with the discovery of the glymphatic system. The process of CSF egress is thought to occur via multiple routes including along the dural lymphatics, perineural pathways, transvenous via AGs, and with additional routes along the spine^2^. The precise proportion of clearance via each route is unclear but likely varies between individuals. Disturbance in this physiologic process which can occur as a result of insults such as SAH may help to understand this system further. The current findings suggest that anatomic variations may dictate the resilience of this system in the setting of pathology. We believe that AGs are important for bulk transvenous egress of CSF. In the setting of SAH, microscopic pathways along the dural lymphatics and perineural routes become obstructed, shifting the clearance more heavily towards the transvenous route via AG. Thus, having a higher number of AG may serve as a protective factor in the development of chronic hydrocephalus. Of course, this is at present only a theoretical consideration and significant further work will be required to elucidate the precise mechanism by which AG provide resilience to this system.

### Limitations

The current study is limited in its retrospective design. As a result, the clinical follow-up for all confounders was not uniformly available. The conclusions which may be drawn are associational and causality remains a question of future work. The sample size utilized for this study is relatively small, however, fairly robust significant results are seen nonetheless. Future work will be required to validate these findings in other retrospective cohorts, as well as using more robust prospective study designs.

## Conclusion

In conclusion, the current study identifies the number of AG as a novel anatomic predictor of shunt dependence. Further work is underway to validate these findings in a different cohort. The findings of this study are of clinical significance and may serve as a radiologic biomarker to predict shunt dependence. The study also provides potential indirect insight into the balance between the routes of CSF egress and will serve as a platform for hypothesis generation.

## Data Availability

All data produced in the present study are available upon reasonable request to the authors

## References

1. Rekate HL. A contemporary definition and classification of hydrocephalus. Semin Pediatr Neurol. 2009;16(1):9–15. doi:10.1016/j.spen.2009.01.002

2. Proulx ST. Cerebrospinal fluid outflow: a review of the historical and contemporary evidence for arachnoid villi, perineural routes, and dural lymphatics. Cell Mol Life Sci. 2021;78(6):2429–2457. doi:10.1007/s00018-020-03706-5

3. Chen S, Luo J, Reis C, Manaenko A, Zhang J. Hydrocephalus after Subarachnoid Hemorrhage: Pathophysiology, Diagnosis, and Treatment. BioMed Research International. doi:https://doi.org/10.1155/2017/8584753

4. Garton T, Keep R, Wilkinson D, et al. Intraventricular Hemorrhage: the Role of Blood Components in Secondary Injury and Hydrocephalus. Translational Stroke Research. 2016;7. doi:10.1007/s12975-016-0480-8

5. Miyajima M, Arai H. Evaluation of the Production and Absorption of Cerebrospinal Fluid. Neurol Med Chir (Tokyo). 2015;55(8):647–656. doi:10.2176/nmc.ra.2015-0003

6. Mondejar V, Patsalides A. The Role of Arachnoid Granulations and the Glymphatic System in the Pathophysiology of Idiopathic Intracranial Hypertension. Curr Neurol Neurosci Rep. 2020;20(7):20. doi:10.1007/s11910-020-01044-4

7. Jessen NA, Munk ASF, Lundgaard I, Nedergaard M. The Glymphatic System: A Beginner’s Guide. Neurochem Res. 2015;40(12):2583–2599. doi:10.1007/s11064-015-1581-6

8. Rasmussen MK, Mestre H, Nedergaard M. The glymphatic pathway in neurological disorders. Lancet Neurol. 2018;17(11):1016–1024. doi:10.1016/S1474-4422(18)30318-1

9. Kanat A, Turkmenoglu O, Aydin M, et al. Toward Changing of the Pathophysiologic Basis of Acute Hydrocephalus After Subarachnoid Hemorrhage: A Preliminary Experimental Study. World neurosurgery. 2012;80. doi:10.1016/j.wneu.2012.12.020

10. Aboul-Ela HM, Salah El-Din AM, Zaater AA, Shehab M, El Shahawy OA. Predictors of shunt-dependent hydrocephalus following aneurysmal subarachnoid hemorrhage: a pilot study in a single Egyptian institute. Egypt J Neurol Psychiatr Neurosurg. 2018;54(1):11. doi:10.1186/s41983-018-0015-1

11. Wilson CD, Safavi-Abbasi S, Sun H, et al. Meta-analysis and systematic review of risk factors for shunt dependency after aneurysmal subarachnoid hemorrhage. Journal of Neurosurgery. 2017;126(2):586–595. doi:10.3171/2015.11.JNS152094

12. Team, R. Core. R: A language and environment for statistical computing; Published online 2018.

13. JASP team. JASP (0.11.1.0).; 2021. https://jasp-stats.org/

14. Ganesh A, Luengo-Fernandez R, Wharton RM, Rothwell PM. Ordinal vs dichotomous analyses of modified Rankin Scale, 5-year outcome, and cost of stroke. Neurology. 2018;91(21):e1951–e1960. doi:10.1212/WNL.0000000000006554

15. Xie Z, Hu X, Zan X, Lin S, Li H, You C. Predictors of Shunt-dependent Hydrocephalus After Aneurysmal Subarachnoid Hemorrhage? A Systematic Review and Meta-Analysis. World Neurosurg. 2017;106:844-860.e6. doi:10.1016/j.wneu.2017.06.119

16. Ramanan M, Lipman J, Shorr A, Shankar A. A meta-analysis of ventriculostomy-associated cerebrospinal fluid infections. BMC Infect Dis. 2015;15:3. doi:10.1186/s12879-014-0712-z

